# Increased aperiodic gamma power in young boys with Fragile X is associated with better language ability

**DOI:** 10.1101/2020.10.08.20209536

**Authors:** Carol L. Wilkinson, Charles A. Nelson

## Abstract

The lack of identified clinical biomarkers in Fragile X Syndrome (FXS), the most common inherited form of intellectual disability, has limited the successful translation of bench-to-bedside therapeutics. While numerous drugs have shown promise in reversing synaptic and behavioral phenotypes in mouse models of FXS, none have demonstrated clinical efficacy in humans. Electroencephalographic (EEG) measures have been identified as candidate biomarkers as EEG recordings of both adults with FXS and mouse models of FXS consistently exhibit increased resting-state gamma power. However, the developmental timing of these EEG differences is not known as thus far EEG studies have not focused on young children with FXS. Further, understanding how EEG differences are associated to core symptoms of FXS is crucial to successful use of EEG as a biomarker, and may improve our understanding of the disorder. Resting-state EEG was collected from FXS boys with full mutation of *Fmr1* (32-84 months old, *n*=11) and compared with both age-matched (*n*=12) and cognitive-matched (*n*=12) typically developing boys. Power spectra (including aperiodic and periodic components) were compared using non-parametric cluster-based permutation testing. Associations between 30-50Hz gamma power and cognitive, language, and behavioral measures were evaluated using Pearson correlation and linear regression with age as a covariate. FXS participants showed increased power in the beta/gamma range (∼25-50Hz) across multiple brain regions. Both a reduction in the aperiodic (1/f) slope and increase in beta/gamma periodic activity contributed to the significant increase in high-frequency power. Increased gamma power, driven by the aperiodic component, was associated with better language ability in the FXS group. No association was observed between gamma power and parent report measures of behavioral challenges, sensory hypersensitivities, or adaptive behaviors. The observed positive association between increased aperiodic gamma power and language supports hypotheses that increased E/I ratios observed in FXS mouse models may reflect beneficial compensation.

## Introduction

Fragile X Syndrome (FXS), an X-linked, single-gene disorder, is the most common inherited form of intellectual disability(1). In addition to cognitive deficits, children with FXS often have significant language impairments as well as behavioral challenges that overlap with several neurodevelopmental disorders including autism spectrum disorder (ASD), attention deficit hyperactivity disorder (ADHD), and anxiety(2). Indeed, virtually all boys with FXS present with some autistic symptoms, and 30-50% meet formal diagnostic criteria for ASD(3,4). Therefore, understanding the neural mechanisms that underlie specific cognitive, language, and behavioral deficits in FXS is both crucial to the development of effective therapeutics, but also may shed light on the pathophysiology of comorbid disorders.

The disorder is caused by an expansion of a CGG trinucleotide repeat on the X chromosome, leading to silencing of the *Fmr1* gene, and reduced expression of its protein product, Fragile X Mental Retardation Protein (FMRP)(2). Although research in rodent models of FXS has improved our understanding of the molecular mechanisms underlying the disorder, there is still no effective treatment for FXS. In addition to mimicking many of the cognitive and behavioral phenotypes seen in humans, the *Fmr1* KO mice have alterations in synaptic and structural plasticity, including impairments in long-term potentiation and depression(5), and alteration in excitatory/inhibitory (E:I) balance(6–9). Whereas several compounds (eg. mGluR5 negative modulators(10), GABA agonists(11,12)) have successfully reversed phenotypes in *Fmr1* KO mice, human phase II trials have disappointingly shown limited effect on outcome measures(13). A lack of brain-based biomarkers in FXS, and other ASD-related disorders has been identified as a major challenge to therapeutic development in the field(13–15).

Electroencephalography (EEG) is a brain-based biomarker candidate, as it is low cost, non-invasive, and has been used in both human and mouse studies of FXS. Further, EEG studies in both FXS adults and *Fmr1* KO mice have observed increased resting-state gamma band power compared to controls, and reduced inter-trial phase synchrony in the gamma band in response to auditory stimuli(16–19). Human studies with FXS individuals have also observed reduced relative alpha power and elevated relative theta power(16,20,21) compared to neurotypical individuals, but this has not been consistently observed in mouse studies.

Aberrant gamma oscillations are particularly intriguing as a brain-based biomarker for FXS and overlapping neuropsychiatric disorders for several reasons. First, gamma oscillations are generated by parvalbumin-expressing inhibitory interneurons, and thereby indirectly represent E:I balance in the cortex(22). Alterations in both inhibitory neurons and gamma oscillations have been observed not only in FXS(23), but several other neuropsychiatric/neurodevelopmental disorders including schizophrenia and ASD(24–29). Second, gamma activity has been associated with clinically relevant processes such as sensory integration, language processing(30–34) and working memory(35,36). Further, alterations observed in the gamma band in *Fmr1* KO mice have been rescued by targeted pharmacological intervention(18).

However, it is still unclear how observed alterations in the gamma band relate to core cognitive and behavioral features of FXS, especially in the developing brain. In a recent, relatively large (*n=*38) EEG study in adults with FXS, increased frontal gamma power was significantly associated with a number of behavioral features reported on the Aberrant Behavioral Checklist (eg. irritability, hyperactivity, stereotyped behaviors), a commonly used parent report measure in clinical trials. In addition gamma power was negatively associated with adaptive measures of communication and a direct measure of cognition. Smaller studies in FXS adults have also found associations between resting-state gamma power and increased sensory sensitivity and social impairment(16). Similarly, in our study of toddlers with familial risk of ASD, we have found increased frontal gamma power to be negatively associated with language development(32), and in boys with ASD increased gamma power has been associated with worse developmental delay(37).

To our knowledge no published studies have characterized baseline EEG activity in young boys with FXS. This is an important gap in the field, as therapeutics (both behavioral and pharmacologic) will ideally be implemented near the age of diagnosis (∼ 3 years of age). In addition, few studies have specifically looked at how differences in brain activity relate to core features of FXS – cognitive and language impairments. Given these gaps, this study had two main goals. First, we aimed to characterize baseline EEG activity in a group of preschool to young school aged boys with full-mutation FXS, as compared to either age-matched or cognitive-matched typically developing boys. We hypothesized that FXS boys would have similar baseline EEG abnormalities observed in FXS adults with increased theta and gamma power. Second, we aimed to characterize the relationship between baseline EEG measures and a range of cognitive measures. This was done in both a hypothesis driven and exploratory manner. We tested two hypotheses based on previous findings: (1) gamma power would be positively associated with various behavioral challenges as measured by the Aberrant Behavior Checklist and (2) gamma power would be negatively associated with cognition and language ability. Given that no prior studies have evaluated EEG and related symptoms in this age range, exploratory analyses investigated possible associations between gamma power and additional parent report measures often used in clinical trials.

## METHODS

### Participants

A total 16 boys (33-78 months old) with full mutation of *Fmr1* and 12 similarly aged (32-80 months old) typically developing boys were recruited for this study (IRB#P00025493) conducted at Boston Children’s Hospital/Harvard Medical School. 4 FXS participants did not complete baseline EEG acquisition (3 net refusal, 1 unable to maintain protocol), and 1 FXS participant’s EEG data was excluded due to excessive artifact. EEG and behavioral data were analyzed for a total of 11 FXS boys (mean age = 53.5 months; SD = 16.4 months) and 12 typically developing boys (mean age = 47.7 months; SD = 13.1).

FXS and age-controls: FXS participants all had documented full mutation of the *Fmr1* gene, but could have size mosaicism (mixture of full and premutation) and methylation status was not known for all participants. Girls were excluded from this study given their variable expression of *Fmr1* and the small size of this study. Across all groups, additional exclusion criteria included history of prematurity (<35 weeks gestational age), low birth weight (<2000gms), known birth trauma, known genetic disorders (other than FXS), unstable seizure disorder, current use of anticonvulsant medication, and uncorrected hearing or vision problems. Some participants were on stable doses of medications (Oxybutin (1 age-matched control); melatonin (2 FXS); Miralax (1 age-matched control). Children were from primarily English-speaking households with English spoken more than 50% of the time (2/11 FXS and 2/12 age-matched control participants were either in bilingual households or daycare).

Cognitive and sex-matched controls: EEG data from an additional set of 12 cognitive-matched boys were analyzed (mean age = 29.8 months, SD 10.1 months, range 14-52 months old). 11 individuals from this group provided EEG data as part of a concurrent longitudinal study (IRB#P00018377) in the lab which used the same EEG resting-state paradigm. Controls were identified by matching FXS participants’ Fine Motor and Visual Reception raw scores on the Mullen Scales of Early Learning (see below). In order to appropriately match all FXS participants, one EEG in this group overlapped with the above age/sex matched control group.

Institutional review board approval was obtained prior to starting the study. Written, informed consent was obtained from all parents or guardians prior to their children’s participant in the study. Table 1 describes participant characteristics.

**Table 1:** Sample Characteristics.

|  | Age-matched<br>N = 12 | FXS<br>N = 11 | Cog-matched<br>N = 12 |
| --- | --- | --- | --- |
| **Age**, mean in months (SD) | 47.6 (13.1) | 53.5 (16.3) | 29.8 (10.1) |
| **Maternal Education, *n* (%)** |  |  |  |
| < 4-year college degree | 0 (0) | 0 (0) | 1 (8) |
| 4-year college degree | 4 (33) | 2 (18) | 2 (17) |
| >4-year college degree | 8(77) | 9 (81) | 9 (75) |
| **Paternal Education, *n* (%)** |  |  |  |
| < 4-year college degree | 0 (0) | 1 (9) | 3 (25) |
| 4-year college degree | 6 (50) | 4 (36) | 6 (50) |
| >4-year college degree | 6 (50) | 6 (54) | 3 (25) |
| **Household income, *n* (%)** |  |  |  |
| <\$40,000 | 0 | 0 | 1 (8) |
| \$40-70,000 | 0 (0) | 3 (27) | 0 (0) |
| \$70-100,000 | 3 (25) | 3 (27) | 0 (0) |
| \$100-140,000 | 5 (42) | 1 (8) | 4 (33) |
| >\$140,000 | 4 (33) | 4 (36) | 7 (58) |
| **Race, *n* (%)** |  |  |  |
| White | 7 (58) | 9 (81) | 9 (75) |
| African American | 0 (0) | 0 (0) | 1 (8) |
| Asian | 1 (8) | 1 (9) | 0 (0) |
| Mixed | 4 (33) | 1 (9) | 2 (17) |
| **Ethnicity, *n* (%)** |  |  |  |
| Hispanic or Latino | 0 (0) | 3 (27) | 1 (8) |
| **24m MSEL NVDQ** Mean±SD | 112.6±15.7 | 58.2±14.3 | 102.8±16.2 |
| Visual Reception Age Equiv<br>(months±SD) | 54.5±8.8 | 32.3±14.4 | 32.2±13.3 |
| Fine Motor Age Equiv (months±SD) | 44.6±7.1 | 29.5±12.0 | 29.5±12.0 |
| **EEG Quality Measures** |  |  |  |
| Number of 2 sec Segments | 116.9±17.4 | 82.1±38.5 | 101.9±34.4 |
| Percent Good Channels | 94.9±4.0 | 94.1±3.6 | 90.6±3.8 |
| Percent ICs Rejected | 36.2±7.6 | 34.0±9.1 | 37.0±10.0 |
| Median Artifact Probability | 0.03±0.02 | 0.07±0.04 | 0.05±0.05 |

### EEG Assessment

Resting-state EEG data were collected in a dimly lit, sound-attenuated, electrically shielded room. The child either sat in their seated caregiver’s lap or sat independently in a chair, high-chair, or stroller based on behavioral preference. Caregivers were instructed by a research assistant to avoid social interactions or speaking with their child. Continuous EEG was recorded for 2-5 minutes depending on compliance. Given known behavioral challenges in the FXS population, parents were asked about expected behavioral challenges, calming techniques, and motivators specific to each child. To improve compliance, participants were shown a silent screensaver of abstract colorful moving images and allowed to hold a fidget toy. EEG data were collected using a 128-channel Hydrocel Geodesic Sensor Net (Version 1, EGI Inc, Eugene, OR) connected to a DC-coupled amplifier (Net Amps 300, EGI Inc, Eugene, OR). Data were sampled at 1000Hz and referenced to a single vertex electrode (Cz), with impedances kept below 100kΩ. Electrooculographic electrodes were removed to improve the child’s comfort.

### EEG pre-processing

Raw NetStation (NetStation version 4.5, EGI Inc, Eugene, OR) files were exported to MATLAB (version R2017a) for pre-processing and power analysis using the Batch EEG Automated Processing Platform (BEAPP; (38)) with integrated Harvard Automated Preprocessing Pipeline for EEG (HAPPE; (39)). Preprocessing has previously been described in detail for similar data(32). Briefly, data were 1Hz high-pass and 100Hz low-pass filtered, downsampled to 250hz, and then run through the HAPPE module for 60Hz line noise removal, bad channel rejection and artifact removal using combined wavelet-enhanced independent component analysis (ICA) and Multiple Artifact Rejection Algorithm (MARA(40,41)). Given the short length of EEG recording, 39 of the 128 channels were selected for ICA/MARA (Standard 10-20 electrodes: 22, 9, 33, 24, 11, 124, 122, 45, 36, 104, 108, 58, 52, 62, 92, 96, 70, 83; Additional electrodes: 23, 28, 19, 4, 3, 117, 13, 112, 37, 55, 87, 41, 47, 46, 103, 98, 102, 75, 67, 77, 72). After artifact removal, channels removed during bad channel rejection were interpolated, data were rereferenced to an average reference, detrended using the signal mean, and segmented into 2-second segments. Any segments with retained artifact were rejected using HAPPE’s amplitude and joint probability criteria.

EEG were rejected for data quality if they had fewer than 20 segments (40 seconds total), or did not meet the following HAPPE data quality output parameters: percent good channels >80%, mean and median retained artifact probability <0.3, percent of independent components rejected <84%, and percent variance after artifact removal <32%. Table 1 shows quality metrics for all groups.

### EEG power analysis

Power spectral density (PSD) at each electrode, for each 2 second segment, was calculated with multitaper spectral analysis(42) embedded in BEAPP using three orthogonal tapers. For each electrode for a given EEG, PSD for each frequency bin (0.5Hz frequency resolution) was averaged across segments, and then averaged across the regions of interest shown in Figure 1. PSDs were normalized using Log10(Hz). Our analyses were limited to absolute power, as relative power measurements were artificially affected by normalization; increased power in high frequency bands in FXS participants artificially lowered the relative power of lower frequency bands. The PSD was also analyzed using the FOOOF v1.0.0 parameterization model across a 2-55Hz frequency range (https://github.com/fooof-tools/fooof; in Python v3.6.8) in order to model periodic and aperiodic components of the power spectra(43). The FOOOF model was used in the fixed mode (no spectral knee) with *peak_width_limits* set to [1, 18.0], *max_n_peaks* = 7, and *peak_threshold* = 2). For each subject’s power spectrum FOOOF provides two parameters to describe the aperiodic 1/f background signal: offset and slope. To determine an aperiodic-adjusted gamma power, the FOOOF estimated aperiodic signal was subtracted from the raw power spectrum, resulting in a flattened spectrum. FOOOF model fit to the original spectrum for ech group is shown in Supplemental Figure 1.

**Figure 1.**
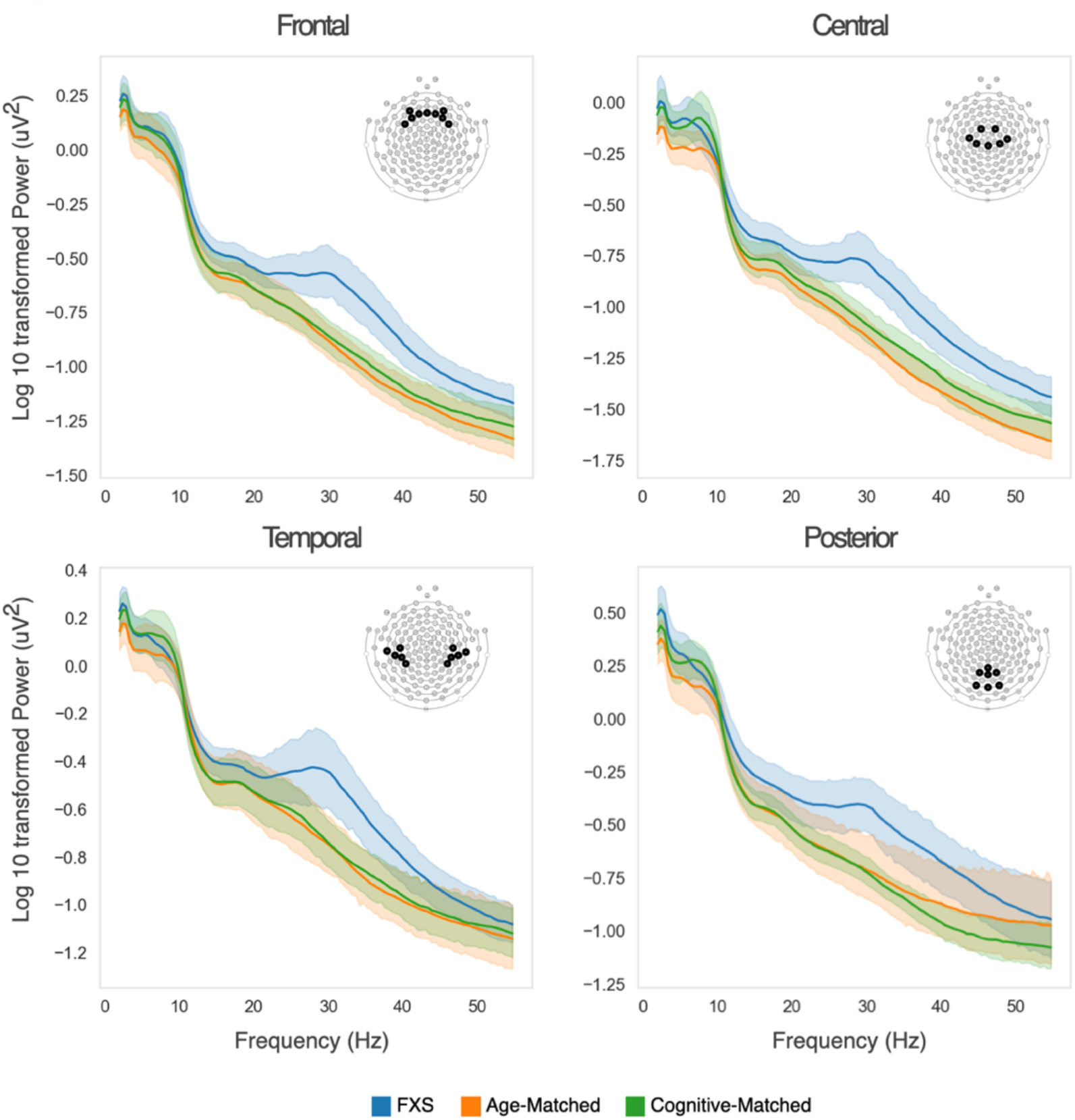
Power spectra across frontal, central, temporal, and posterior regions of interest. Log10 transformed power spectra of FXS (blue), age-matched controls (green), and cognitive-matched controls (orange) are shown for each region of interest. Shaded areas describe 95% confidence intervals.

### Behavioral Measures

The Mullen Scales of Early Learning (MSEL(44)) is a standardized cognitive measure for children 0-69 months of age. Non-verbal subscales (fine motor, visual reception) were administered to all FXS participants regardless of age, age-matched controls under 70 months of age, and all cognitive matched controls, and the Nonverbal Developmental Quotient (NVDQ) was calculated. The Preschool Language Scale 5^th^ Edition (PLS(45)), a comprehensive developmental language assessment standardized for children 0-83 months of age, was administered to FXS and age-matched participants. Standard scores of subtests for receptive (Auditory Comprehension) and expressive (Expressive Communication) language, as well as the total standard score were calculated. Note that per research administration protocol augmentative communication devices are not used during this assessment, so scores may underestimate a child’s non-verbal expressive language skills. The following clinical questionnaires were completed by primary caregivers of FXS and age-matched participants: Aberrant Behavior Checklist-Community Edition(46) (ABC-FXS, scored using FXS specific factoring system(47)), Vineland Adaptive Behavior Scales, Third Edition(VABS-3(48), Repetitive Behavior Scale-Revised(49), and Sensory Profile, Child-2(50). The ABC-FXS scoring included 6 subscales: irritability, hyperactivity, lethargy, social avoidance, stereotypy, and inappropriate speech.

### Statistical Analyses

T-test, or Mann Whitney if data was not normal in distribution, was used to compare differences in behavioral scores or EEG measures between either FXS *vs*. age-matched controls, or FXS *vs*. cognitive-matched controls. To examine group differences in the power spectra, a non-parametric clustering method, controlling for multiple comparisons using Monte Carlo estimation (1000 permutations)(51) was employed with MNE-Python(52) using a F-statistic threshold of 4.32.

Regression analysis was used to characterize the relationship between gamma power and behavioral measures within the FXS group. Analysis were performed used Stata software, version 14.2 (Stata). Figures were created using Python v3.6.8 and python data visualization libraries (*matplotlib*(53) and *Seaborn* (https://seaborn.pydata.org/index.html).

## RESULTS

### Sample Description

Demographic data, including the MSEL non-verbal developmental quotient (NVDQ) and subtest age-equivalents are shown in Table 1. As expected, FXS participants had significantly lower nonverbal skills compared to age-matched controls (t-tests; MSEL NDVQ: p < 0.0001, Fine Motor: p < 0.01, Visual Reception: p<0.001), but had similar age-equivalent scores to the cognitive-matched control group.

Groups were also similar on EEG quality metrics and well below our quality thresholds (see methods). However the average number of EEG segments available for power analyses was significantly lower in the FXS group compared to age-matched (Mann Whitney, p<0.05), but not cognitive-matched control groups (Mann Whitney, p>0.05).

### Absolute Power Spectra

Power spectra for all three participant groups across frontal, temporal, central, and posterior regions of interest are shown in Figure 1. FXS power spectra differences were most prominent in the beta-gamma range compared to both age-matched controls and cognitive-matched controls across all regions of interest. A non-parametric clustering method, controlling for multiple comparisons, was used to identify significant differences in the power spectra (Table 2). Analysis of the frontal power spectra, identified a cluster between 23-55Hz for FXS vs. age-matched controls (p=0.004), and between 22-50Hz for FXS vs cognitive-match controls (p=0.007) with significant group differences. Similar clusters were identified for the central power spectra. In contrast, clusters identified for temporal power spectra were smaller in range (26-40Hz), with p-values at, or just below the critical alpha (0.05). For posterior power spectra, the cluster identified between FXS and age-match controls was not significant, however between FXS and cognitively-match controls a cluster between 19-47Hz was identified (p=0.014). Several lower frequency clusters were also identified however none of these were significantly different between groups.

**Table 2:** Power spectra clusters with significant group differences.

| Region of Interest | FXS vs Age-Matched |  | FXS vs Cognitive-Matched |  |
| --- | --- | --- | --- | --- |
|  | Frequency Range (Hz) | Cluster p-value | Frequency Range (Hz) | Cluster p-value |
| Frontal | **22.9-54.7** | **0.004** | **22.0-50.3** | **0.007** |
| Frontal | 12.7-15.6 | 0.188 | - | - |
| Central | **19.0-54.7** | **0.003** | **21.5-52.2** | **0.009** |
| Central | 2.9-5.9 | 0.196 | - | - |
| Central | 12.2-18.6 | 0.132 | - | - |
| Temporal | **25.9-40.0** | **0.041** | 26.4-39.6 | 0.053 |
| Posterior | 23-55 | 0.073 | 11.7-15.6 | 0.147 |
| Posterior | - | - | **22-50** | **0.014** |

Visually, the power spectra of FXS participants also appear to have a reduced slope, suggesting that both aperiodic and oscillatory activity are altered. Growing evidence suggests that aperiodic activity (defined as 1/f^x^, with x = slope of the aperiodic curve) in part represents broad neural firing and balance of excitation and inhibition(54). Further aperiodic activity changes with age and with behavior(55,56), underscoring its likely functional role in cognition. To better understand differences in the aperiodic and oscillatory power spectra components in FXS participants, we used the Fitting Oscillations and One-Over-F (FOOOF) algorithm(43) to estimate aperiodic 1/f (Figure 2A) and periodic/oscillatory components (Figure 2B) of the power spectra. While there were no significant differences in the offset of the aperiodic power spectrum or the aperiodic slope in the FXS group (Figure 2C,D), the aperiodic slope was consistently reduced in the FXS group across frontal, central, and temporal regions of interest.

**Figure 2.**
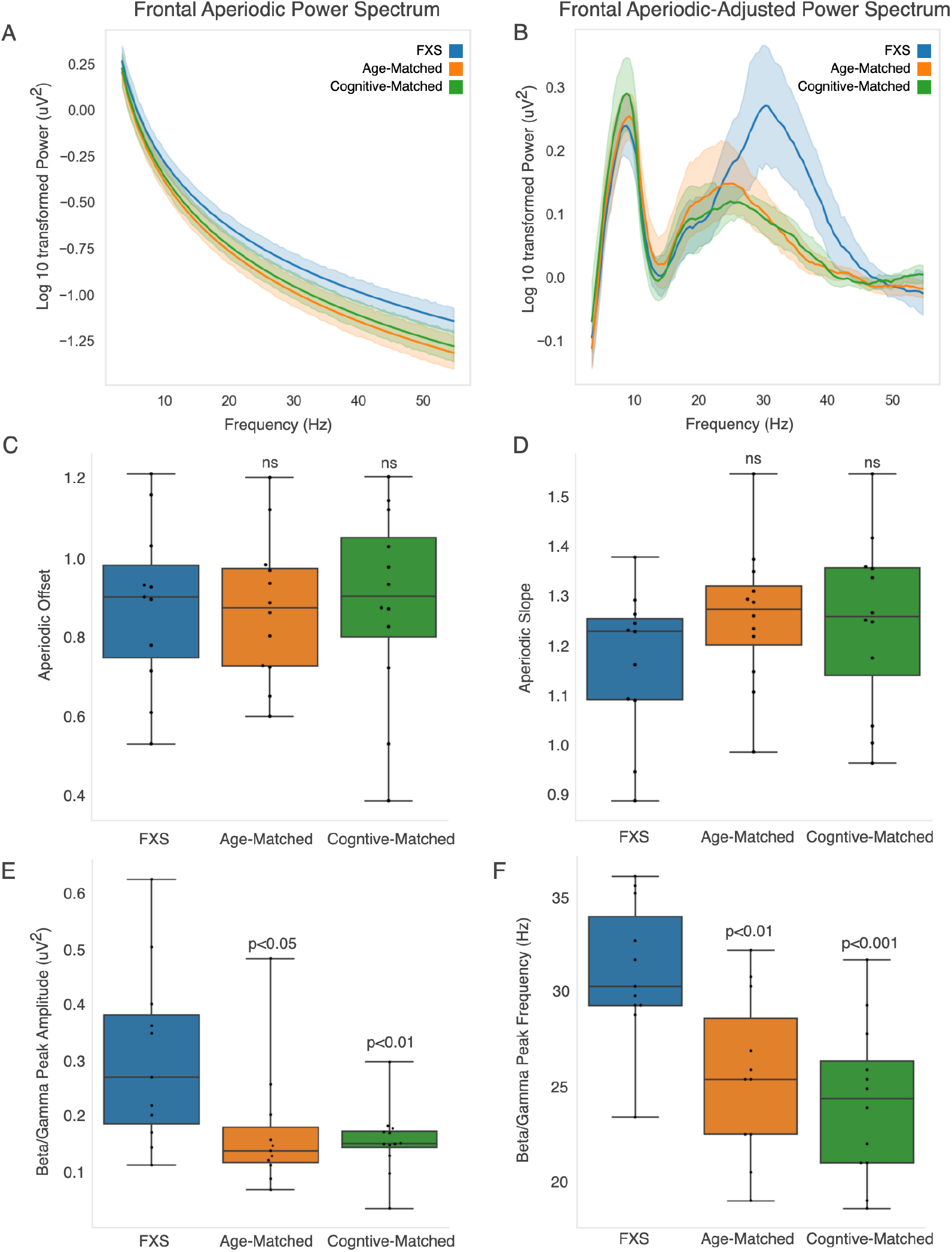
**A:** FOOOF estimated aperiodic power spectrum FXS (blue), age-matched controls (green), and cognitive-matched controls (orange). **B:** FOOOF estimated aperiodic power spectrum was subtracted from the raw power spectrum to determine aperiodic-adjusted power spectrum. Shaded areas describe 95% confidence intervals. **C:** FOOOF estimated aperiodic offset. **D:** FOOOF estimated aperiodic exponent. **E:** Amplitude of maximal peak from Aperiodic-Adjusted Power Spectra between 15-40Hz. **F:** Frequency of identified maximal peak. P-values represent t-test comparisons with FXS group.

Permutation cluster analysis of the frontal aperiodic-adjusted power spectra identified a cluster between 28-41Hz for FXS vs. age-matched controls (p=0.003), and between 26-43Hz for FXS vs. cognitive-match controls (p=0.005) with significant group differences. Similar significant clusters were identified for the other regions of interest. Further analysis of this frequency range found both a significant shift in frequency peak frequency and significant increase in peak amplitude for the FXS group compared to age-matched and cognitive matched controls (Figure 2E, F).

### Frontal Gamma Power and Associations with Clinical Outcomes

Given the above findings, and previous studies in adults with FXS demonstrating significant increase in frontal gamma power, we focus our following analyses on this frequency band alone. Power in the gamma frequency band was calculated in two ways. First, PSD calculations between 30-50Hz were averaged to determine average gamma power for each group (Figure 3A). FXS participants had significantly higher gamma power (−0.87±0.16uV) compared to age-matched (−1.12±0.16 uV; p =0.001) and cognitive-matched (−1.08±0.14 uV; p=0.003) controls. Given group differences in the number of EEG segments analyzed, we confirmed that gamma power was not associated with number of EEG segments retained. Second, we calculated aperiodic and periodic components of gamma power, using the FOOOF estimated aperiodic and aperiodic-adjusted power spectra shown in Figure 2A,B. Both the aperiodic and aperiodic-adjusted gamma were significantly elevated in FXS participants compared to age- and cognitive-matched comparison groups (Figure 3B, C).

**Figure 3.**
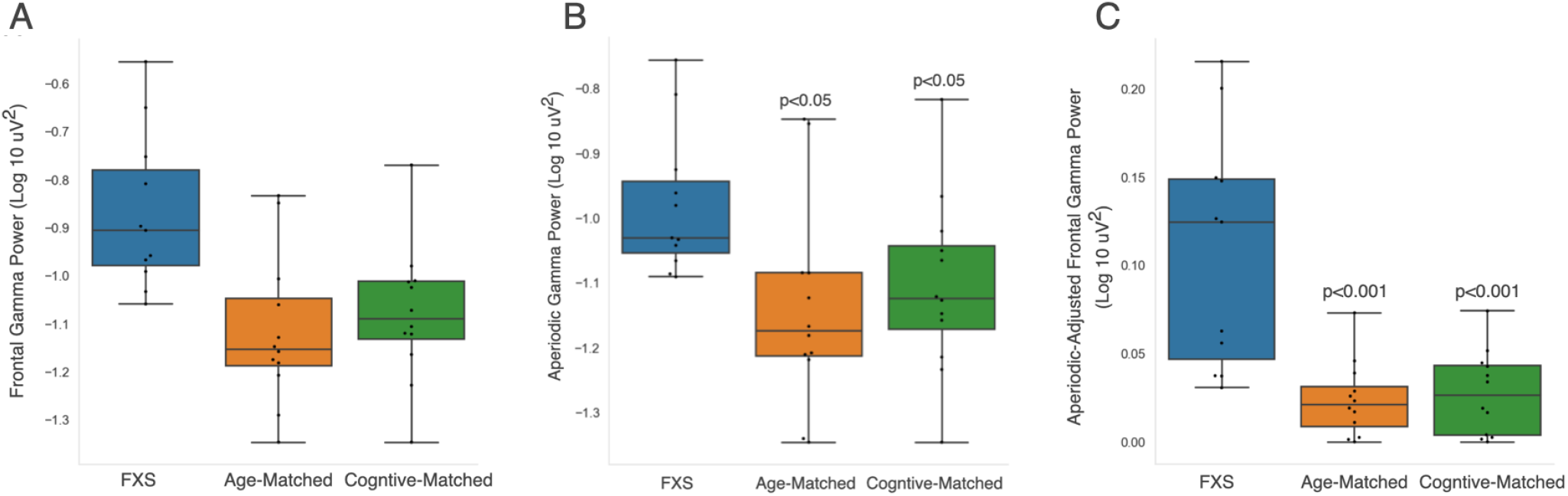
Power in the gamma frequency band (30-50Hz) derived from either the (A) frontal power spectra shown in Figure 1 A, (B) aperiodic frontal power spectra shown in Figure 2A, or (C) aperiodic-adjusted frontal power spectra shown in Figure 2B. P-values represent t-test comparisons with the FXS group.

We next investigated whether increased frontal gamma power in FXS participants was associated with three clinical measures: (1) parent report on the Aberrant Behavior Checklist (ABC-FXS), (2) non-verbal cognitive skills measured by the Mullen Scales of Early Learning (MSEL) nonverbal developmental quotient, and (3) language development as measured by the Preschool Language Scale-5 (PLS-5) as well as parent report on the Vineland Adaptive Behavior Scale (VABS-3, Communication subscale).

There was no significant correlation between frontal gamma power and scores on any of the six ABC-FXS subscales (Pearson R range: −0.31 to 0.17), or with the MSEL nonverbal developmental quotient (Pearson R: 0.16). In contrast, there was a significant and unexpected *positive* correlation between frontal gamma power and standard scores on language measures (Figure 4) both based on behavioral assessment (PLS-5; Pearson R = 0.75; p = 0.007) and parent report (VABS-3, Communication Subscale; Pearson R = 0.62 p = 0.04). Increased gamma power was associated with *better* language skills. There was no observed relationship between frontal gamma power and language in the age-matched comparison group. Given the large age range of participants, and the possibility that age may influence both standard scores and gamma power, a linear regression analyses were performed with age (in months) included as a covariate (Table 3). The positive association between gamma power and language ability as measured by the PLS-5, but not the VABS-3, remained significant after adjusting for age.

**Figure 4.**
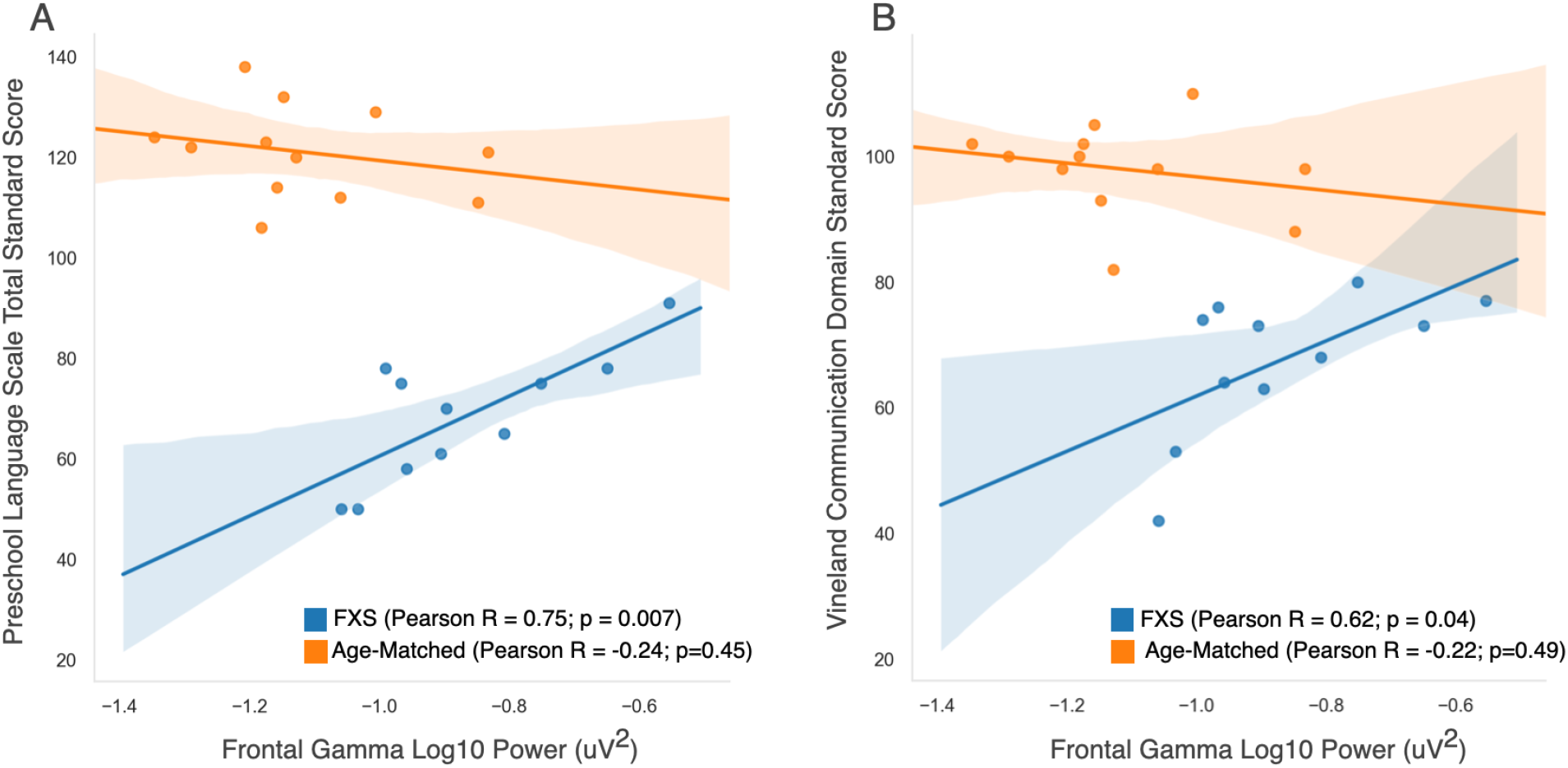
Correlation graphs between overall frontal gamma power (30-50Hz) and (A) total standard score on the Preschool Language Scale; and (B) standard score on the communication domain of the Vineland Adaptive Behavior Scale.

**Table 3:** Linear regression models of gamma power and language measures.

| Dependent Variable | Preschool Language Scale |  | VABS Communication Subscale |  |
| --- | --- | --- | --- | --- |
|  | Model 1 | Model 2 | Model 1 | Model 2 |
| Adjusted R<sup>2</sup> | 0.52 | 0.47 | 0.32 | 0.34 |
| Variables [B Coefficient (SE)] |  |  |  |  |
| Intercept | 120.2 (15.2) | 120.3 (16.1) | 105.8 (16.3) | 105.1 (16.0) |
| Frontal gamma power | **59.6 (17.3)\*\*** | **64.5 (23.6)\*** | **43.9 (18.5)\*** | 26.1 (23.4) |
| Age (months) | - | 0.08 (.23) | - | -0.28 (.23) |

\* = $p < 0.05$
\*\* = $p < 0.01$

To further dissect the functional relevance of increased aperiodic and periodic gamma activity in FXS participants, we also investigated the relationship between aperiodic and periodic/oscillatory components (labeled aperiodic-adjusted) of the gamma band and these two language measures. Notably, the association between gamma and PLS-5 Total Standard Score appears to be driven by the aperiodic component of the gamma band (Figure 5). The adjusted R^2^ was low in models using the aperiodic-adjusted gamma (Table 4), whereas models using the aperiodic component of gamma had high adjusted R^2^ values, and the positive association between aperiodic gamma and the PLS-5 scores remained significant when age was included in the model. While there was a significant positive correlation between aperiodic-adjusted gamma and the VABS-3 Communication Subscale standard score, this did not survive adjustment for age. Post-hoc analyses revealed that age was highly correlated with aperiodic-adjusted gamma (Pearson R = −0.80; p= 0.003) but was not correlated with either aperiodic gamma band activity (Pearson R = −0.37; p=0.27) or with aperiodic slope (Pearson R = 0.16) (Supplemental Figure 4).

**Figure 5.**
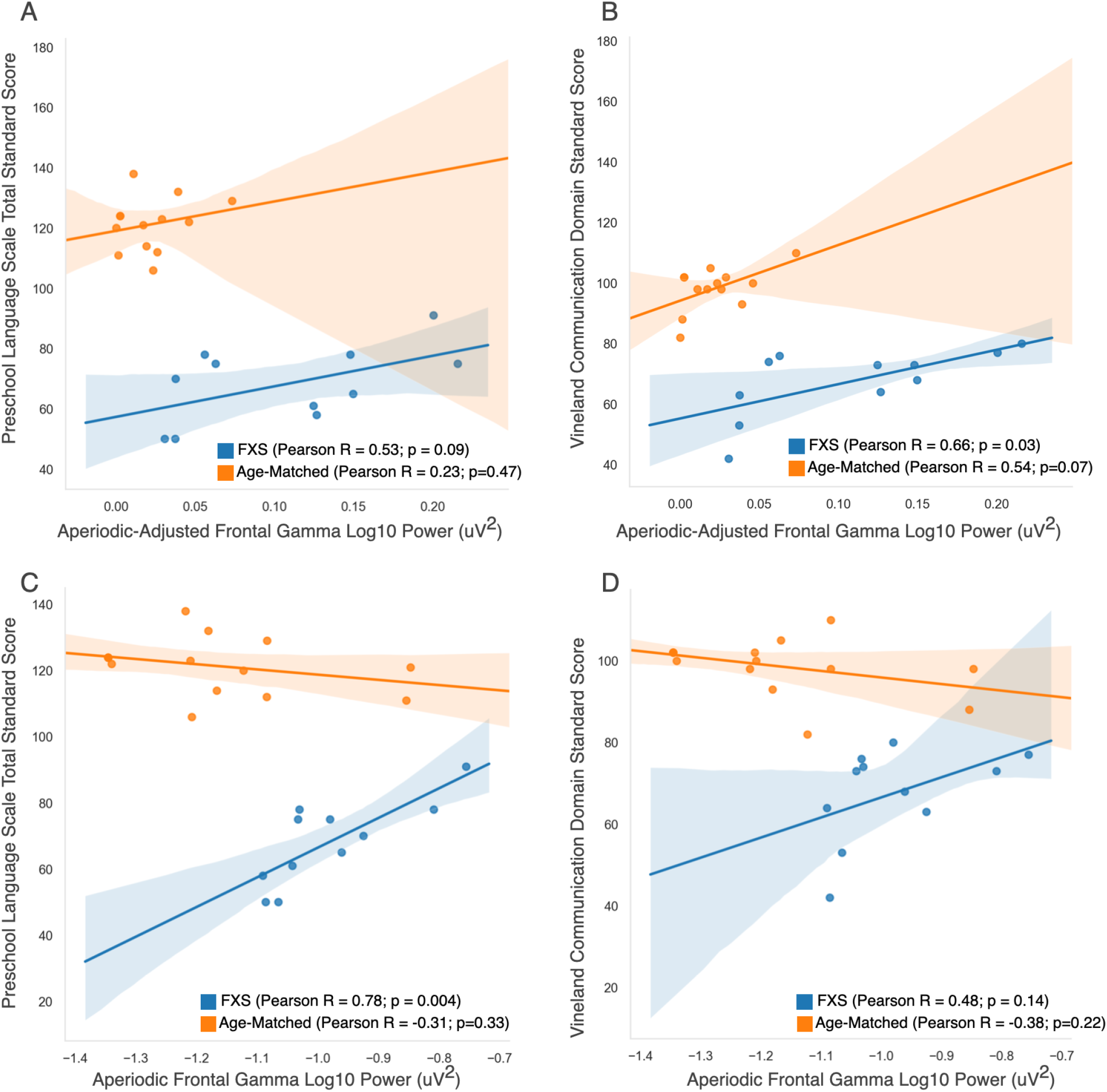
**Top (A,B):** Correlation graphs between aperiodic-adjusted frontal gamma power and language measures. **Bottom (C, D):** Correlation graphs between aperiodic frontal gamma power and language measures.

**Table 4:** Linear Regression FOOOF estimated gamma power measures and language.

|  | **Preschool Language Scale** |  | **VABS Communication Subscale** |  |
| --- | --- | --- | --- | --- |
|  | Model 1 | Model 2 | Model 1 | Model 2 |
| Adjusted R<sup>2</sup> | 0.20 | 0.1 | 0.37 | 0.33 |
| Variables [B Coefficient (SE)] |  |  |  |  |
| Intercept | 57.3 (6.8) | 56.4 (30.3) | 55.3 (5.4) | 69.8 (23.4) |
| **Aperiodic-Adjusted Gamma** | 101.4 (54.0) | 103.7 (96.3) | **113.5 (42.8)\*** | 75.4 (74.4) |
| Age (months) | - | 0.1 (0.39) | - | -0.19 (0.3) |
|  | Model 1 | Model 2 | Model 1 | Model 2 |
| Adjusted R<sup>2</sup> | 0.56 | 0.53 | 0.14 | 0.33 |
| Variables [B Coefficient (SE)] |  |  |  |  |
| Intercept | 156.3 (23.8) | 156.3 (24.6) | 115.8 (29.9) | 115.8 (26.3) |
| **Aperiodic Gamma** | **89.8(24.2)\*\*** | **83.2 (26.8)\*** | 49.2 (30.3) | 29.1 (28.8) |
| Age (months) | - | -0.12 (0.18) | - | -0.37(0.20) |

Additional exploratory analyses of clinical correlations found no significant associations with parent report measures of sensory hypersensitivity as measured by the Sensory Profile-2, other adaptive subscales on the VABS-3 (eg.daily living, social skills, motor skills), or repetitive behaviors as reported on the Repetitive Behavior Scale.

## Discussion

In this study we compare resting (non-task related) EEG power spectra of young boys with FXS to both age-matched and cognitive-matched typically developing boys. Consistent with previous studies in both adolescents and adults with FXS, as well as mouse models of FXS, we observed an increase in power from roughly 25-50Hz across multiple brain regions. Previous studies in adults have also reported increases in resting-state theta power. In this study, permutation cluster analysis identified a single cluster crossing the theta band in the central electrodes, however this cluster did not meet statistical significance. Further, we observed that the increase in high-frequency power is the result of both a decrease in the slope of the aperiodic signal and an increase in high-frequency (∼25-40Hz) activity. Surprisingly, within the FXS group, increased high-frequency activity (as measured by the gamma band 30-50Hz) was associated with *better* language ability.

While the sample size of this study is small, it is the first EEG study focused on preschool to young school aged boys with FXS. Acquiring high quality EEG data from this younger population is challenging, however it is crucial to understanding how underlying neuropathophysiology in FXS changes across development, and to identifying biomarkers that reflect specific core impairments. Notably, identified differences between FXS power spectra and both age-matched and cognitive matched comparison groups were very similar, suggesting that these differences are specific to the genetic disorder, and not due to differences in age or cognitive ability.

### Aperiodic Power Spectra Findings

The measured power spectrum from EEG is the result of a mixture of signals including an aperiodic, broadband background signal, and narrowband oscillatory components. Here we observe that FXS participants have a reduced aperiodic slope, but similar aperiodic offset, leading to overall significantly increased aperiodic activity in the higher frequency bands. Computational modeling suggests that changes in the aperiodic slope are in part driven by the balance between excitation (E) and inhibition (I), with an increased E:I ratio leading to a reduced slope(54). This is consistent with hyperexcitable states observed in *Fmr1* KO mice(6,8,57), and evidence of inhibitory dysfunction in FXS(6,7,23,58). Further, elegant experiments by Antoine et. al.(9) observed increased E:I ratios in the primary somatosensory cortex of four ASD-mouse models, including *Fmr1* KO mice. Interestingly, increased E:I ratios were *not* accompanied by expected increased firing rates or increased postsynaptic potentials in layer 2/3 neurons. Indeed, *in vivo* recordings from awake *Fmr1* KO mice showed no changes in spontaneous firing of regular spiking layer 2/3 neurons and reduced whisker-evoked firing. The authors hypothesize the observed increased E:I ratio represents compensation for altered cortical spiking in order to normalize firing rates. This raises the question of whether increased E:I ratios are functionally beneficial to individuals with FXS. Our language findings support this hypothesis.

Based on previous studies in adults with FXS, we originally hypothesized that increases in gamma power in FXS participants would be negatively associated with language development. However, if increased aperiodic gamma activity reflects homeostatic compensation for ongoing altered cortical spiking resulting in an E:I imbalance, but normalized firing rates, then we may expect improved language development in individuals with the most appropriate homeostatic compensation, especially at a young age. Here we demonstrate that gamma power, specifically the aperiodic signal in the gamma range, was positively associated with language development. Further, model fit was quite strong in our linear regression models limited to aperiodic gamma with and without age as a covariate, with more than 50% of the variance of PLS-5 language scores explained. However, model fit was notably worse for predicting scores on the VABS-3 Communication Subscale, and associations between gamma power and VABS-3 scores were not significant once age was included in the model. The VABS-3 is a parent report measure that focuses more on functional communication skills that can also be impacted by additional factors such as social interest and anxiety. Therefore high-frequency aperiodic activity may specifically be relevant to language development. Indeed, aperiodic gamma power was not associated with non-verbal cognitive skills as measured on the MSEL, or parent report of attention, irritability, sensory hypersensitivity or repetitive/stereotyped behaviors.

The specificity of the association between aperiodic gamma activity and language, instead of for example overall cognition is perhaps surprising, given the growing view that aperiodic or broadband gamma activity reflects overall levels of cortical activity, rather than responses to specific sensory stimuli. Based on this study, we hypothesize that a reduced aperiodic slope, or increased aperiodic gamma, may be associated with improved gamma-phase locking during language or auditory tasks specifically for the preschool age. This is in contrast to what has been observed in FXS adult and *Fmr1* KO EEG studies, where increased background gamma activity is associated with reduced inter-trial phase synchrony in the gamma band in response to a modulated auditory “chirp” stimulus(59,60). However, such associations may change over development; Wen and colleagues have found enhanced evoked gamma responses in juvenile (P21) and adult (P60) *Fmr1* KO mice, but reduced responses at P30. Additional studies are needed to evaluate the relationship between aperiodic gamma and evoked gamma in this younger age range.

### Aperiodic-Adjusted Power Spectra Findings

We also observed increased power between 25-40Hz in the periodic component of the power spectra for FXS participants, with an oscillatory peak observed in most individual tracings (Supplemental Figure 2) at the border of the canonical beta/gamma frequency bands (∼30Hz). This peak was both significantly higher in amplitude and shifted from 25 to 30Hz compared to both age- and cognitive-matched participants. Aperiodic-adjusted gamma overall was not strongly associated with language measures.

This may reflect several variables. First, in our study sample aperiodic-adjusted gamma was strongly negatively correlated with age, making associations with other variables more difficult given our sample size. The negative association between gamma and age has previously been observed in this age range(37). Larger studies in this young age range are needed to better understand both the developmental course of EEG measures in this population, and their relation to core symptoms. Second, this analysis did not include EEG collected as part of a language or auditory based task. Without a stimulus input, it is unclear what narrowband resting-state beta/gamma activity represents.

Mechanistically, benzodiazepines (GABAA receptor modulators) increase EEG measured beta activity, especially in the upper range (21-30Hz), and this effect in rats is even further increased when animals are sitting or walking(61). Increases in EEG measured beta and gamma power have also been associated with muscle artifact(62), however when this occurs measure increases are usually topographically specific to frontal or temporal regions. Here, the shift in beta/gamma peak and increased amplitude was similarly observed across all brain regions (Supplemental Figure 3). Interestingly, robust increases in spontaneous frontocentral beta oscillations (peaking at 23Hz) have also been observed in children with Dup15q syndrome(63). Further investigation of what mechanistically drives ∼30Hz oscillations may shed further light on the pathophysiology of FXS.

### Limitations and Future Directions

The sample size of this study is small. Recruitment of rare genetic disorders is challenging and ultimately large studies will require collaboration and coordination across multiple sites. Further, collecting EEG data in young children with FXS can be challenging as they have significant behavioral challenges including sensitivity to be touched (especially on their heads), limited expressive language, difficulty sitting in one place, reduced attention, and challenges following directions. This study demonstrates that while EEG acquisition in this age group is feasible, 12/16 (75%) participants cooperated with EEG net placement, success requires a research team experienced in both EEG acquisition and behavioral management of challenging behaviors.

Given the above behavioral challenges, consistency in behavior across participants during EEG acquisition was reduced. While most age-matched typically developing children watched the screen saver during baseline EEG acquisition, FXS participants’ attention to the screen was reduced and sometimes required redirection or use of other non-social distractors. It is possible that differences between groups and between individuals are related to this variability in behaviors and emotional states. We do note that our cognitive-matched group was composed of largely toddler-aged boys (14-37 months old), who we expect to have age-appropriate challenges with attention and behavior. In addition, EEG quality metrics were similar across groups. Finally, pre-clinical mouse studies have observed increased gamma power in *Fmr1* KO mice during periods of no movement(17), suggesting that these findings now observed in children, adults, and mice with FXS are reliable and robust.

## Conclusions

Baseline EEG measures from this study in preschool age boys similarly identified gamma abnormalities previous documented in adults and mouse models. However, the unexpected relationship of these EEG abnormalities to clinical symptoms, highlights the complexity of neurodevelopmental disorders such as FXS and the need for larger studies across early childhood in order to further understand changes in EEG measures across development and how these measures relate to clinical symptoms and outcomes.

## Data Availability

The data sets used and/or analyzed during the current study are available from the corresponding author upon reasonable request.

## Acknowledgements

We thank all the families who participated in this study. We also thank Jack Keller, Megan Lauzé, John Fitzgerald, Megan Hartney and the Translational Neuroscience Center Human Neurobehavioral Core at Boston Children’s for their assistance in data collection.

## Supplemental Figures located at the end of the document

### Ethics approval and consent to participate

Institutional review board approval was obtained from Boston Children’s Hospital (IRB#P00025493, IRB#P00018377) prior to starting the study. Written, informed consent was obtained from all caregivers prior to their children’s participation in the study.

### Author Contributions

CLW performed study conception and design, EEG data acquisition, performed the EEG and behavioral data analysis, interpreted the data, and drafted the manuscript. CAN was involved in study conception and design, overseeing data acquisition, and critically reviewed the manuscript for intellectual content.

### Consent for publication

Not applicable.

---

**Supplemental Figure 1.**
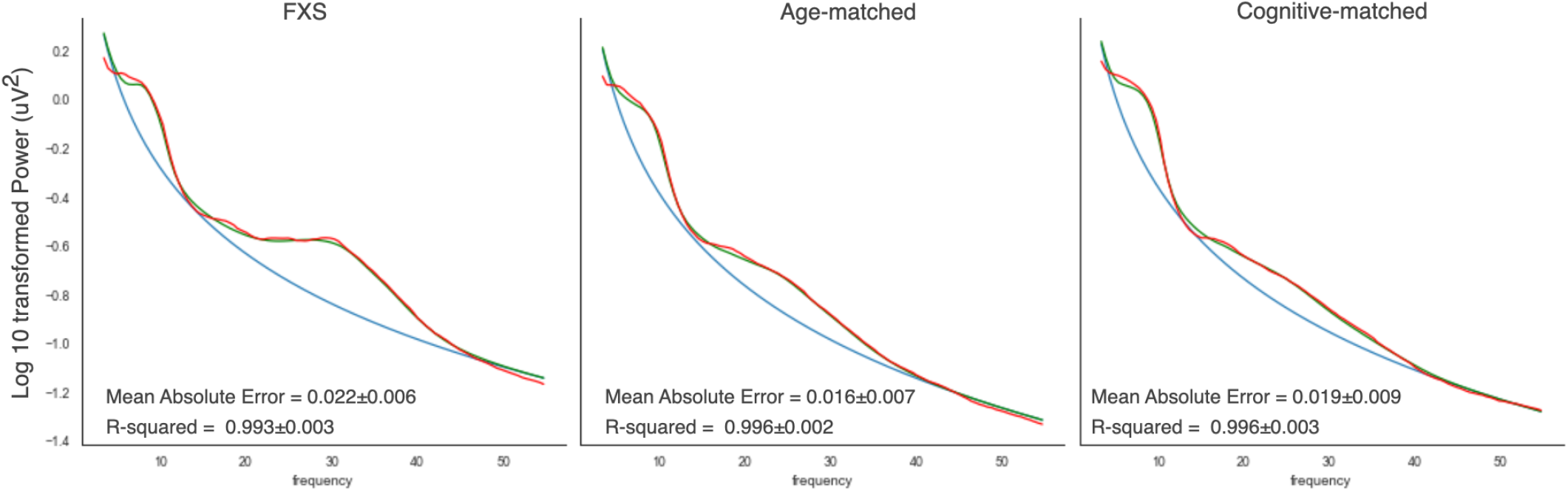
FOOOF parameterization of power spectra. FOOOF estimated aperiodic (blue) and periodic (red) components are shown for each group. Actual averaged power spectra shown in green.

**Supplemental Figure 2.**
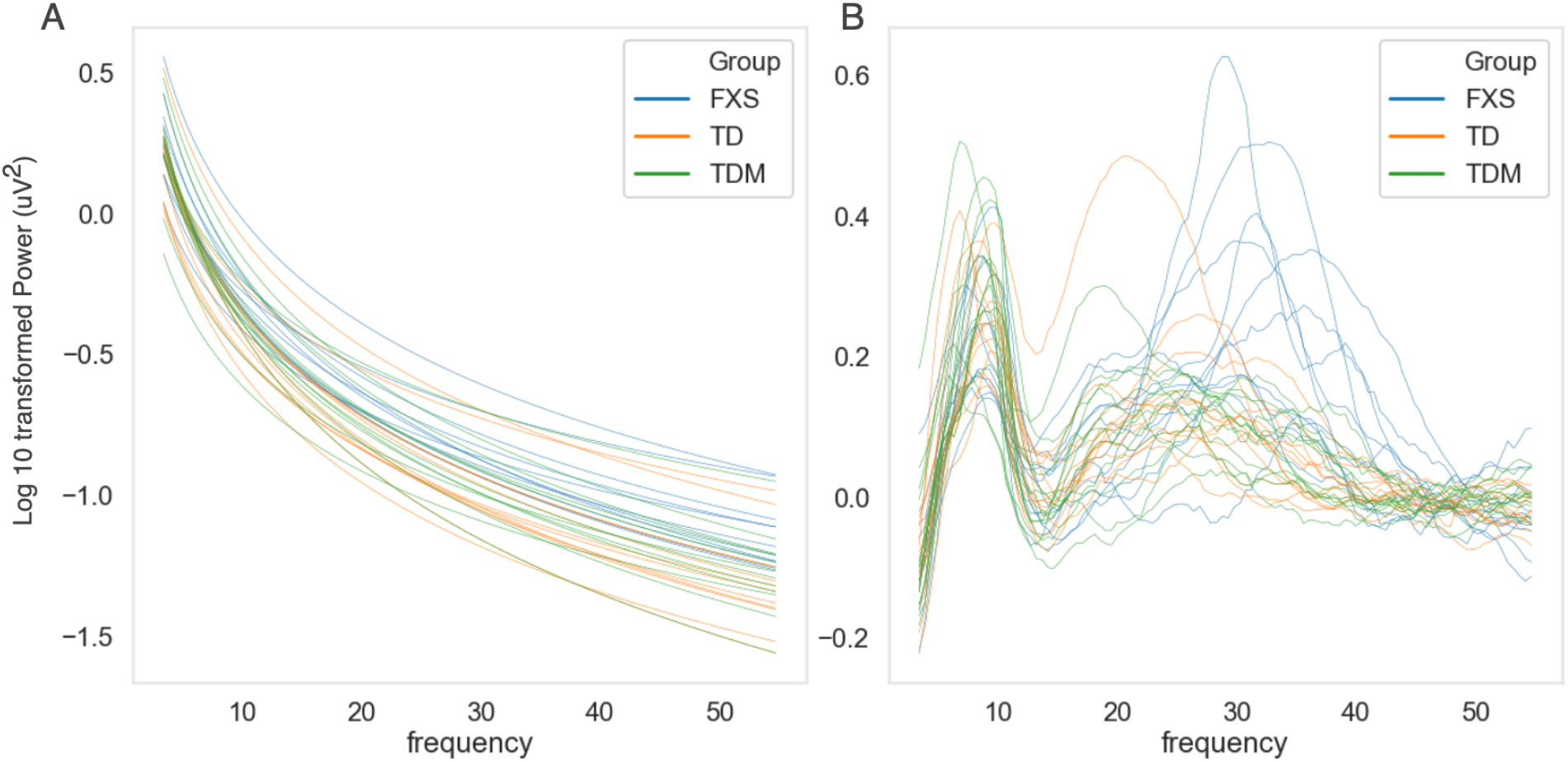
Individual Plots of (A) Aperiodic and (B) Aperiodic-Adjusted power spectra.

**Supplemental Figure 3.**
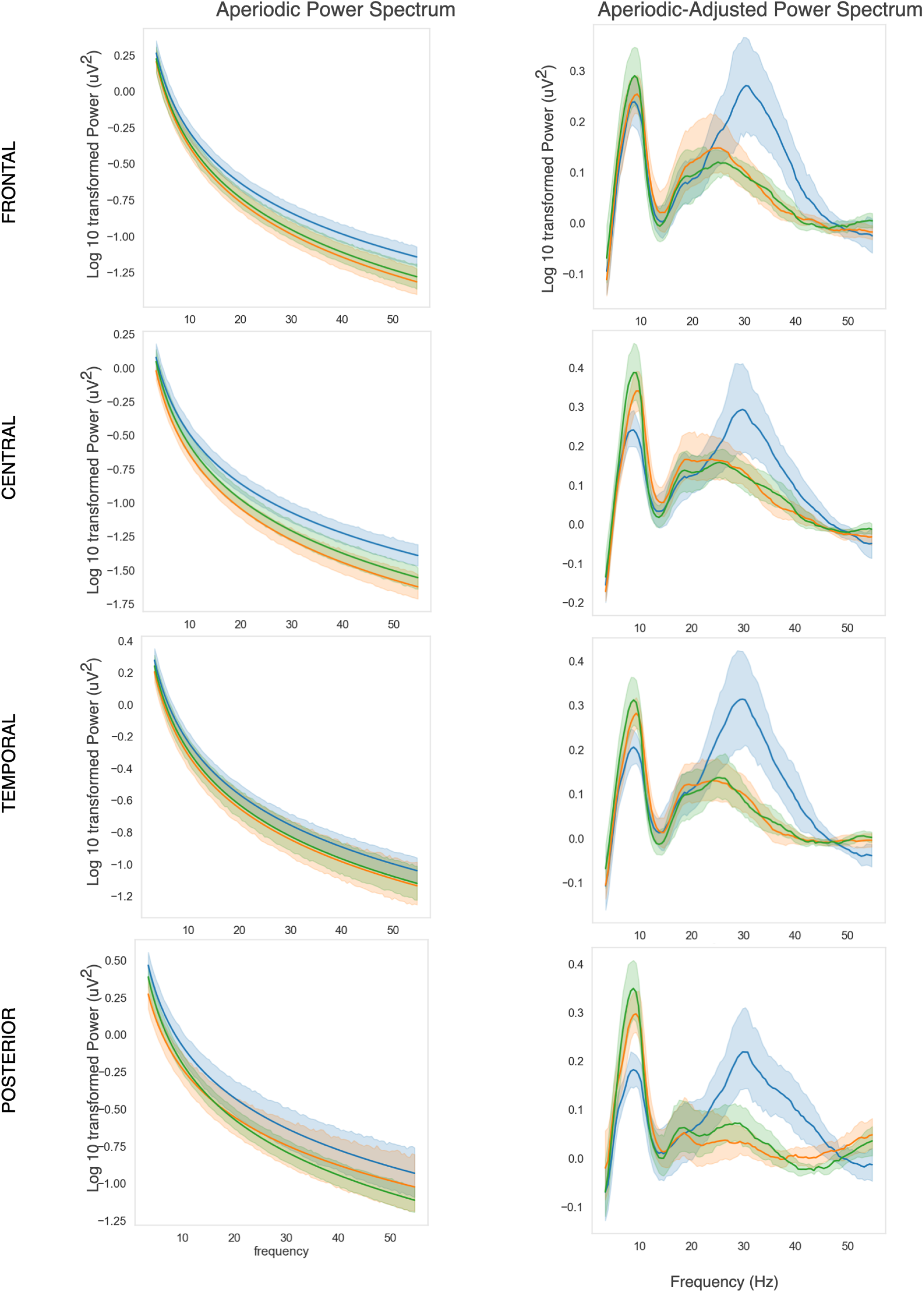
Plots of Aperiodic and Aperiodic-Adjusted power spectra for all regions of interest.

**Supplemental Figure 4.**
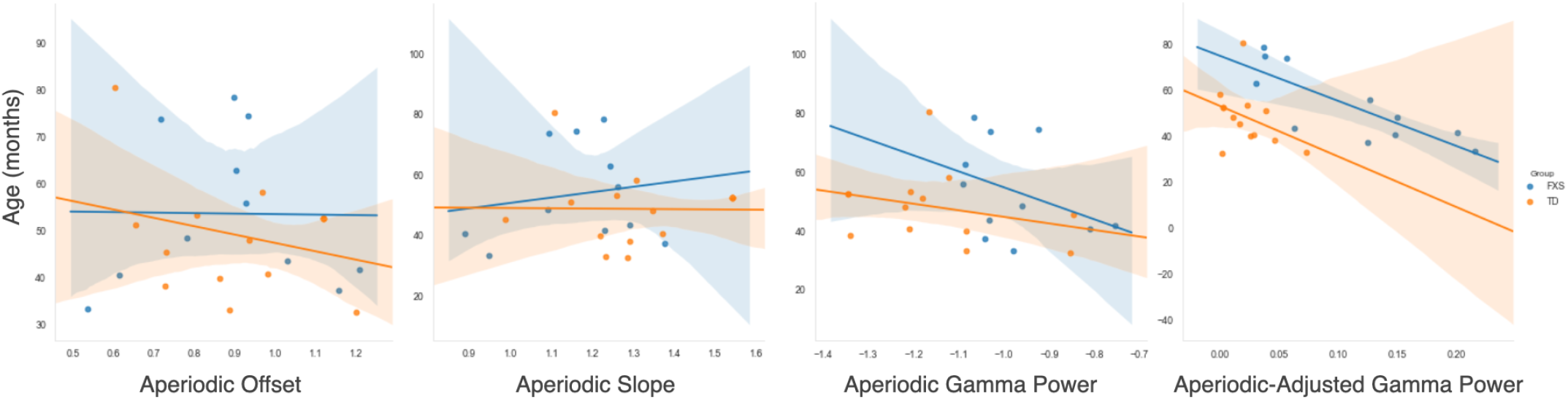
Association of Age and power spectra measures.

